# Developing a functional precision medicine assay to predict pathological complete response in patients with triple-negative breast cancer: Interim results from the PEAR-TNBC Trial

**DOI:** 10.1101/2024.10.25.24314885

**Authors:** Peter Hall, Matthew Williams, Eleonora Peerani, Elli Tham, Francesco Iori, George Richard Tiger Bevan de Fraine, Kerrie Loughrey, Andreas Dimitri Kaffa, Thomas David Laurent Richardson, Carolina Liberal, Angeliki Velentza-Almpani, Demi Annemarie Wiskerke, Farah Sangkolah, Aston Martin Crawley, Jay Kearney, Nourdine Kabirou Bah, Marios Konstantinos Tasoulis, Cliona C Kirwan, Susan Cleator, Steve Chan, Duleek Ranatunga

## Abstract

**Introduction:** More than 150,000 women die worldwide every year of Triple-Negative Breast Cancer (TNBC). There are a range of treatment options, but no good way to match patients to their optimal treatment. For most newly diagnosed patients with early TNBC, the current standard of care is neoadjuvant chemo/immunotherapy before surgery, with patients who achieve a pathological complete response (pCR) having a better prognosis.

We have developed a Functional Precision Medicine (FPM) test that uses a fresh biopsy, dissociates the cells, embeds them in a 3D hydrogel matrix, cultures them in a microfluidics device and tests them against a range of systemic therapies, while using a computer vision pipeline to measure responses to therapies ex vivo.

**Methods:** We designed and conducted an observational multi-centre clinical trial to assess the feasibility of using our FPM assay in patients with newly diagnosed TNBC undergoing neoadjuvant therapy. Patients underwent an additional core needle biopsy followed by systemic therapy as part of routine care. We assessed the response in our assay against whether patients achieved pCR or not at the time of definitive surgery, and calculated Receiver-Operating Characteristic curves (ROC) to optimize cut-offs. In patients who did not achieve pCR, we explored whether there were other regimens that had a better in-assay performance.

**Results:** In cohort A, we recruited 34 patients, of whom 12 are evaluable as of 31st July 2024 All were female. Nine patients achieved a pCR. Specificity was 100%, sensitivity 78%, p = 0.0455 and the AUC for the ROC for predicting pCR vs. non-pCR was 0.78. In the 3 patients who did not achieve a pCR, one patient had a regimen that performed better in assay than the treatment they received, and where the response was greater than the cut-off that predicted pCR in other patients.

**Conclusion:** We have presented interim results from a novel FPM assay in patients with early stage TNBC. Our test demonstrates good performance in predicting pCR. The trial continues to accrue data, and Cohort B continues to recruit (PEAR-TNBC; NCT05435352).

**CoI statement:** Williams, Peerani, Tham, Iori, de Fraine, Loughrey, Kaffa, Richardson, Liberal, Velentza-Almpani, Wiskerke, Sangkolah, Crawley, Kearney, Bah, Ranatunga are employees of Pear Bio, with salary, stock options and IP.

Hall has received honoraria from Pfizer, Eisai, MSD, Seagen, Exact Sciences, Gilead, AstraZeneca and conference expenses from Lilly and Novartis.

Williams has research funding or agreements from Cancer Research UK, Breast Cancer Now, The Brain Tumour Charity, Brain Tumour Research and Novocure.

Tasoulis has received honoraria from the BMJ and Company: BMJ and IntegraConnect

Kirwan reports no Conflict of Interest

## Background

Precision medicine approaches have transformed the management of many cancers, particularly in diseases such as EGFR-mutated lung cancer, and HER2 positive breast cancer. However, there are two problems with such approaches. Firstly, biomarkers are an imperfect predictor of response: 20% of patients with EGFR mutated lung cancer do not respond to EGFR-targeted agents (1), and even with dual HER2 targeting and chemotherapy, only 80% of patients with HER2 positive breast cancer achieves an objective response (2). Secondly, most cancers, and the majority of those causing cancer deaths, lack actionable biomarkers (3,4).

An alternative approach is to directly assess tumor response to treatment. This is technically demanding, and has been explored using a range of approaches and platforms. In general, they require isolation of live cells from fresh tissue samples, and those samples being directly exposed to potential drugs to assess their impact. Previous approaches have included organoid-based approaches and stem cell assays (5,6). These functional models measure direct response of a patient sample in an *ex vivo* model, and the use of functional models to guide therapy is referred to as Functional Precision Medicine (FPM).

However, validating such approaches is challenging. As well as the technical challenges around isolating and culturing tumor cells, which may vary across tumor types, there are challenges in interpreting different responses from 2D vs. 3D vs. *in vivo* xenografts, and how these relate to responses in patients (e.g. response vs. survival). As a result, there are no FPM assays in routine clinical use.

Breast cancer is the most common cancer in women, and is responsible for more than 11,000 deaths per year in the UK and 42,000 in the USA. Although many breast tumors express estrogen, progesterone and HER2 receptors, approximately 15% of primary breast cancers do not express any of these three markers - “triple negative” breast cancer (7). Such tumors do not respond to either hormone-based or HER2 targeted agents, and so the mainstay of medical treatment is cytotoxic chemotherapy and immunotherapy. Although various regimens are used, there is no good data comparing different commonly-used regimens, and patterns of practice vary.

The diagnosis of breast cancer is typically made on a combination of clinical examination and imaging, with pathological confirmation. However, in newly diagnosed early TNBC, the use of neoadjuvant chemotherapy/immunotherapy is now standard-of-care, and the rate of complete response, as assessed by pathologist (pCR) has consistently been shown to predict patient outcomes (8). The risk of recurrence and death is reduced by 76% and 81%, respectively, in patients who achieve a pCR following neoadjuvant therapy compared to those who do not (9). Neoadjuvant chemotherapy/immunotherapy for TNBC is therefore well suited to FPM approaches: there are a range of treatment options, with no clear way to decide between them, with a clear binary outcome (pCR vs. no pCR) and where that outcome has significant impact on long-term patient survival. Although there is now good data from randomized trials that show the improvement in pCR with newer regimens, such trials measure an average, not a per-patient, effect and inter-patient variation in response to therapy remains challenging to predict, and intensified regimens carry a higher risk of toxicity (both medical and financial). Therefore, being able to predict individualized benefit would allow a better understanding of the trade-off between benefits and side-effects.

Developing new predictive tests requires us to measure their sensitivity (true positive rate) and specificity (true negative rate). For a test where we have a continuous predictive variable and a binary outcome we need to decide what level of test result provides the best cut-off point between “positive” and “negative” cases (10), accepting that no single cut-off point will perfectly distinguish positive and negative cases. This is most often done by plotting the true positive rate (sensitivity) against the false negative rate (1-specificity) as we vary the threshold we use to decide between “positive” and “negative” cases. As we collect data on more cases, this plot begins to resemble a curve - the Receiver-Operating Characteristic (ROC) curve. The area under the curve provides a measure of test performance and the shape of the curve can help decide where to place the threshold. Once we have decided on a threshold, we can then use a 2x2 grid (a confusion matrix) to display predicted and actual positive and negative patient numbers, and plot the per-patient result in relation to the threshold chosen to show how sensitive the results are likely to be to changes in threshold in a separation plot.

We have developed a platform that uses fresh tumor-extracted cells embedded in a 3D hydrogel matrix to test systemic therapies side-by-side in an *ex vivo* functional precision medicine assay. Microtumors are encapsulated and imaged daily on days 0-3 using confocal microscopy. Each microtumor is housed in a supportive chip, which contains connection points for drug perfusion and clearance. Two chips are kept as a negative control (media only), while the others are treated with a range of different systemic therapy agents, including combinations. Computer vision is used to analyze the 3D confocal micrographs to allow counting of live/dead cell number, cell viability and culture width over time. These measures are then correlated with outcomes seen in clinical practice.

In this paper we present the initial results from the PEAR-TNBC Cohort A clinical study which provides early data to support the validation of a functional precision medicine assay in TNBC. We also discuss some of the challenges in delivering a functional precision medicine assay.

## Methods

### Dose finding

*Ex vivo* doses for use in the Pear Bio assay were established through a combination of 2D and 3D cell line assays followed by testing on 3D primary patient microtumors. Briefly, 3 breast cancer cell lines (MBA-MB-157 (ATCC, HTB-24), MBA-MB-231 (ATCC, HTB-26), MDA-MB-468 (ATCC, HTB-132)), were used to obtain dose-response curves of drugs as monotherapies using an Incucyte^®^ SX5 System (Sartorius). IC50 and IC75 values were obtained from the dose response curves. Combination therapies were empirically validated using a 10x10 dose grid and analyzed using a Bliss model (11). IC50 doses, and 10x higher and lower doses were tested on cell lines encapsulated in our breast cancer hydrogel formulation for 3D cell culture (BREA-F49) using RealTimeGlo(R) assay (Promega, G9711). Doses that showed efficacy across 2D and 3D cell assays for each drug were taken forward for testing on primary cells in 3D using the Pear Bio assay setup. Pembrolizumab drug dose was determined using flow cytometry of PBMCs to assess PD1 expression and proliferation. Final doses selected for the assay are reported in Appendix 1.

### 3D tumor cell culture

We formulated a bespoke hydrogel formulation, BREA-F49 using a combination of PureCol™ EZ Gel solution (Sigma-Aldrich, 5074), Matrigel^®^ (Corning, 354230) and Sodium Hyaluronate (Lifecore Biomedical, HA40K-5).

In order to further characterize our models, we conducted RNA sequencing to analyze the transcriptional profile of the cells prior to and after encapsulation within our hydrogel formulation. The transcriptional profiles of tumor-extracted cells grown in 3D in BREA-F49 for 4 days were compared to cell pellets from the same patient (N=3). Briefly, tumor cells were extracted as described below, split into 2 aliquots, one to be stored in triplicates as pellets, one to be encapsulated in BREA-F49 at 1,000 cells/µL in 100 µL in triplicates and cultured for 4 days. Hydrogels were dissociated in TRIzol^TM^ (ThermoFisher, 15596026) followed by a Qiagen RNeasy Micro Kit (Qiagen, 74004), alongside thawed cell pellets, for RNA extraction following manufacturer’s instructions. Sequencing was performed by the Imperial College BRC Genomics Facility as polyA RNAseq with 30M reads, paired-end 75 bp on a Nextseq2000 P3 200cyc machine.

### Patient sample processing

Patient samples were placed in 6mL of tissue storage medium (T-Store Tissue Storage and Transportation Medium, Life Science Production) and transported to Pear Bio’s central lab at 4°C on the same day of biopsy (day -1). Core needle biopsies were washed over a 100 uM strainer before being weighed. Samples were transferred into Omics digestion pouches and were dissociated into a mixed single cell suspension via enzymatic digestion (Tumor Dissociation Kit human, Miltenyi Biotec + Y-27632 (STEMCELL Technologies, 72304)) using an optimized tissue-specific program on the Cytiva VIA Extractor. Post-dissociation, the mixed single cell suspension was strained through a 70 uM strainer, before being washed and centrifuged and the cell pellet then resuspended and counted using a acridine orange/propidium iodide cell viability dye, read on the LUNA-FL™ Dual Fluorescence Cell Counter (Logos Biosystems)) to quantify total, live, dead cell populations and viability.

Up to 120,000 cells were taken, resuspended in BREA-49 at a 1 million cells/μL density and then plated into equal aliquots of 10μL containing 10,000 cells per aliquot. 2 aliquots were placed in each microfluidic chip. Where we were able to form > 6 aliquots, we used two aliquots to test the same treatment, creating duplicates. Each aliquot was embedded in a 10μL of liquid BREA-F49, and left to crosslink for 45 minutes in a humidified incubator at 37°C (this is referred to as the core). After crosslinking, a further 80μL of PureCol™ EZ Gel solution (Sigma-Aldrich, 5074) was added to fill the microfluidic chip chamber and crosslinked in an incubator for 45 minutes. The microfluidic chip was then sealed, filled with cell culture medium and placed overnight in a humidified incubator (Fig App3.2). Chips receiving the Keynote-522 regimen had 25μg/ml of pembrolizumab-containing cell culture medium added to each chip prior to overnight incubation. The next day was defined as Day 0 (baseline), at which point we conducted baseline microscope imaging, then connected each chip to a pump and microfluidic device system which enabled the sequential flow and clearance of therapeutic regimens.

Each chip was imaged daily on days 0-3, prior to returning each chip in its spot in the microfluidics device. Cells were stained daily prior to imaging for one hour with 200nM MitoView 633 (70055, Biotium) and 2μM NucView 488 (30029, Biotium). Imaging was performed using a Leica Stellaris i5 confocal microscope with 10x air objective lens. Images were obtained at a resolution of 512 x 512 pixels, acquiring images at 5μm interval vertically (z axis step size) through the whole depth of gels. Digital images were transferred to cloud storage, and analyzed using internally-developed computer vision (CV) software.

Cells were classified as being dead if they were seen in the NucView 488 channel, dying if dual-stained with MitoView and Nucview and alive if they were only seen in the Mitoview 633 channel. Cell viability was calculated as live cells divided by total cells (Live + Dead - Dying). Cell migration (in μm/day) was measured using single particle tracking (12) implemented in the CV pipeline using the Laptrack package (13). We measured the total numbers of cells, live cells, dying cells and dead cells and mean, median, top 5% and maximum speed for migration, and plotted these daily. We assessed culture viability by monitoring changes in live cell counts over time in control microtumors, stipulating as a quality control measure that the live cell count in the negative control (untreated) microtumors on the final day of microscope imaging (day 3) should be ≥70% of the live tumor cells counted at baseline microscope imaging (day 0) for the sample to be considered evaluable.

Each microtumor was treated with a range of different systemic therapy agents, starting after imaging on day 0 (baseline). Drugs are made up at the required concentration with primary culture medium in a 15mL falcon tube reservoir. This is hooked to the peristaltic pump and the drugs are perfused into each chip as per their drug treatment condition using the doses determined using the methods described above. Microtumors were imaged daily for days 0-3, and CV software was used to count and classify cells as above. For patients where we had enough cells to establish 2 microtumors per treatment, we averaged all metrics across both microtumors.

To assess the response to therapy, we looked at changes in the number of dead cells. We calculated the “dead cell delta” by calculating the change between the dead cell count on Day 3 and Day 0; we then normalized that by dividing the difference by the dead cell count at Day 0 to yield a “normalized dead cell delta” (NDCD) metric to account for the fact that not all microtumors started with exactly the same number of cells.

Microtumors were treated with different therapy regimens (Table 1), based on trial data an international guidelines (14–16) From patient 7 onwards pembrolizumab was added to the therapy list based on Keynote-522 trial results (17)

**Table 1.** Regimens used in the Pear Bio test with order of administration and trial references. All regimens used the same dose of each drug across different conditions. The clearance period consisted of a change of drug reservoirs and any excess drugs in each microfluidic chip removed manually. Details of doses and suppliers are provided in Appendix 1.

| Acronym | Regimen description | Drugs and timings used in the Pear Bio assay | Trial reference |
| --- | --- | --- | --- |
| AC-T | Doxorubicin and cyclophosphamide followed by paclitaxel | Doxorubicin (24 hours)<br>Cyclophosphamide (24 hours) (Clearance)<br>Paclitaxel (48 hours) | Early Breast Cancer Trialists Group (14) |
| AC-Carbotaxol | Doxorubicin and cyclophosphamide followed by carboplatin and paclitaxel | Doxorubicin (24 hours)<br>Cyclophosphamide (24 hours) (Clearance)<br>Carboplatin (48 hours)<br>Paclitaxel (48 hours) | ESMO guidelines (15) |
| T-AC | Paclitaxel followed by doxorubicin and cyclophosphamide | Paclitaxel (48 hours) (Clearance)<br>Doxorubicin (24 hours)<br>Cyclophosphamide (24 hours) | NCCN guidelines (16) |
| Carbotaxol-AC | Carboplatin and paclitaxel followed by doxorubicin and cyclophosphamide | Carboplatin (48 hours)<br>Paclitaxel (48 hours) (Clearance)<br>Doxorubicin (24 hours)<br>Cyclophosphamide (24 hours) | ESMO guidelines (15) |
| Pembro-Carbotaxol | Carboplatin, paclitaxel and pembrolizumab followed by | Pembrolizumab (48 hours)<br>Carboplatin (48 hours) | Keynote-522 (17) |
| /Pembro-AC/<br>Pembro | doxorubicin,<br>cyclophosphamide and<br>pembrolizumab | Paclitaxel (48 hours)<br>(Clearance)<br>Doxorubicin (24 hours)<br>Cyclophosphamide (24 hours)<br>Pembrolizumab (24 hours) |  |

### Clinical Trial

The PEAR-TNBC study (NCT05435352) was open to newly diagnosed patients with a histological diagnosis of TNBC. Patients had to be aged 18+, with stage 1-3 TNBC where the primary tumor was ≥10mm and could not have had any previous treatment. Patients with inflammatory or metastatic cancer were not eligible. Patients were enrolled from 5 tertiary oncology centers in the UK.

All patients provided written informed consent to participate in the study, and the study obtained relevant ethical and research approvals (London Queen Square REC: 21/PR/1027). At enrolment, patients underwent a core needle biopsy procedure and provided two 14 gauge core needle biopsy samples, which were placed in T-store medium and processed as described above.

The primary aim was to assess the specificity of the Pear Bio assay in predicting pCR vs non-pCR in patients with early-stage TNBC receiving neoadjuvant therapy. Secondary aims included the sensitivity and positive and negative predictive value of the Pear Bio assay in predicting pCR. Exploratory aims included an analysis of culture success rates, subgroup analysis and the relationship between biomarkers, response in the Pear Bio assay and patient outcomes.

Patients underwent neoadjuvant therapy and surgery as per local standard of care for TNBC. Post-operative samples were reviewed by local pathology and classified as pathological complete response (ypT0/is ypN0) (18). Patients who did not achieve a pCR were further classified as having partial response, stable disease or disease progression based on local investigator assessment. For the purposes of the interim analysis, we considered outcomes only as pCR vs. non-pCR. Patients were considered evaluable if we had managed to establish microtumors and test the treatment they received alongside a non-treated control for 3 days using a microscopy readout, and if the patient had undergone at least 4 cycles of neoadjuvant therapy followed by surgery. Safety monitoring was limited to post-biopsy complications for the 3 days post-biopsy. The data was locked for analysis on 31st July 2024.

Actual response was based on the pathological result at surgery, categorized as pCR or non-pCR. Treating clinicians were blind to the results of the assay. We used the normalized dead cell delta (NDCD) to predict response to therapy. For evaluable patients, we plotted the magnitude of NDCD against actual response. We optimized the threshold of the metric by constructing a threshold plot and inspecting the range of values for patients achieving a pCR and not, constructing a receiver-operating characteristic (ROC) curve and calculating the area under the curve (AUC) of the ROC. Once we had identified the best threshold, we used that to predict which patients were likely to achieve a pCR or not. Since therapy choice was independent of the results of the assay, we constructed a confusion matrix of predicted vs. actual response and used to calculate sensitivity, specificity, accuracy and positive and negative predictive values. We calculated a p-value for the contingency table using Fisher’s exact test given the small sample size. In patients who did not achieve a pCR, we examined whether there were alternative regimens that were predicted to lead to a pCR.

We summarized predicted therapy effectiveness by considering the rank of each therapy (the treatment with the highest predicted efficacy being rank 1). We looked for evidence of patterns in ranking by visualizing the ranks in a stacked donut plot, and calculating Kendall’s W (where 1 equals perfect agreement in rank order between different metrics and 0 denotes no agreement). We explored whether there were one or two regimens that were persistently best by calculating how many patients had samples that followed each rank ordering.

## Results

IC50 values were determined in three TNBC cell lines using normalized confluency readouts at either 24 hours (doxorubicin, cyclophosphamide) or 48 hours (carboplatin, paclitaxel) on an Incucyte SX5 system (Sartorius) (Figure App2.1). Doses were confirmed in 3D by running the Real-Time-Glo assay on cell lines cultured in BREA-F49 hydrogel, where no major differences in responses was observed. These values were taken forward as doses to be used in PEAR-TNBC.

Since treatment regimens in neoadjuvant TNBC include combination therapies, we tested chemotherapies in combination in order to assess potential synergistic or antagonistic effects between the drugs *in vitro*. Combination therapies were empirically validated using a 10x10 dose grid and analyzed using a Bliss model (19). No strong antagonistic or synergistic drug effects were observed (Figure App2.2).

RNA sequencing was conducted on breast cancer cell pellets pre and post culture in our 3D hydrogel formulation BREA-F49 (day 3 of culture, N=3). Retention of immune cells in tumor-extracted cell pellets pre and post encapsulation and culture in our BREA-F49 hydrogels was confirmed through cell deconvolution analysis (N=3). Fig App2.3 shows immune cells were retained following dissociation (in tumor-extracted cell pellets) and also retained in comparable percentages throughout 3D cell culture (gel). Retention of tumor-resident immune cells in our cultures is key to successful testing of immunotherapies (e.g. pembrolizumab) using this system.

### Trial Results

The trial opened to recruitment on 27th May 2022 and completed recruitment from 5 sites to Cohort A on 4th July 2024. We approached 66 patients, of whom 36 gave informed consent and 34 participated (consent rate of 55%). At the time of analysis, 12 patients were evaluable. Patients received a range of treatments as part of standard clinical care. Patient demographics, treatment and disease response are shown in Table 2, and patient data flow in Figure 2.

**Figure 1:**
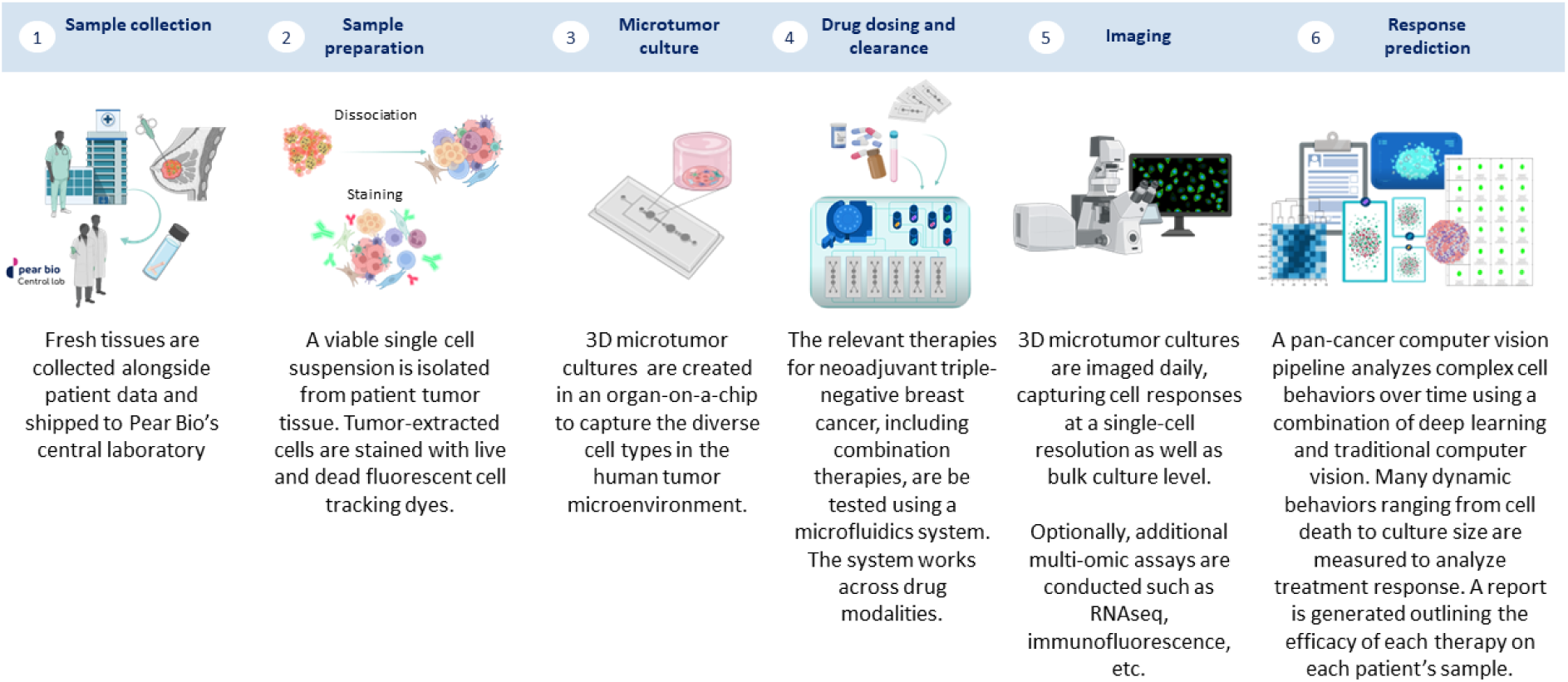
Patient sample collection and analysis pathway.

**Figure 2:**
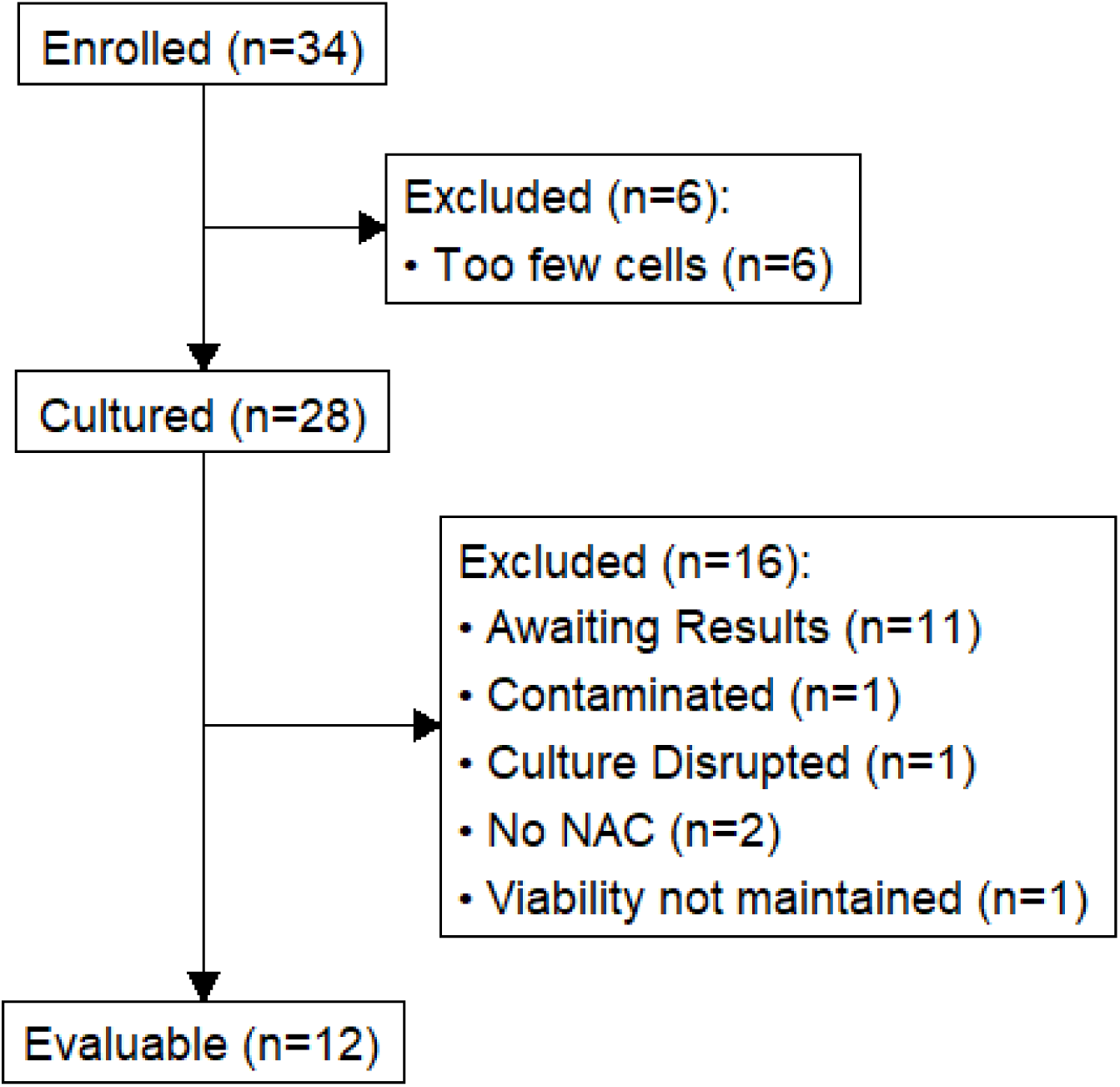
CONSORT diagram for PEAR-TNBC trial. Twelve patients were evaluable at the time of the interim analysis (data lock 31^st^ July 2024). NAC = neoadjuvant chemotherapy.

**Table 2:** Patient and treatment details for all evaluable patients. Treatment types are those received by patients as part of routine care.

|  |  |
| --- | --- |
|  | Evaluable Patients (n=12) |
| Age (Median, IQR) | 53 (47-60) |
| Sex | All female |
| Ethnicity |  |
| Asian | 4 |
| White | 5 |
| Other | 1 |
| Missing | 2 |
| Stage | Pre-treatment |
| Stage 1 | 3 |
| Stage 2A | 4 |
| Stage 2B | 1 |
| Stage 3A | 1 |
| Stage X | 3 |
| T Stage |  |
| T0 | 0 |
| Tis | 0 |
| T1 | 2 |
| T2 | 7 |
| T3 | 1 |
| Tx | 2 |
| N Stage |  |
| N0 | 8 |

|  |  |
| --- | --- |
| N1 | 1 |
| Nx | 3 |
| Tumour size in mm (Median, IQR) | 27 (25-35) |
| Treatment | Keynote-522: 7 |
|  | AC-CarboTaxol: 4 |
|  | EC-CarboTaxol: 1 |
| Median number of cycles ( range) | 8.5 (4-16) |
| pCR | 9 (75%) |

Nine patients achieved a pCR. Commonly used treatment regimens were AC-CarboTaxol and Keynote-522. Seven patients received Keynote-522. Per-patient staging, treatment and outcomes are presented in Table 2. No patients developed complications from their biopsy, and at time of reporting, no patients have developed recurrent/metastatic disease, and all are alive.

### Patient Assay data

The median number of cells isolated per patient was 552,000 (IQR: 321,000 - 986,000), with a median viability of 66% (IQR: 56-76%); 6 patients had > 300 000 cells. Live cell count in the control microtumors was greater than 70% of starting culture live cell count at day 3 for all patients (Figure App4.1). We plotted changes in NDCD daily for each patient against each treatment regimen (Figure 3 & App4.2).

**Figure 3:**
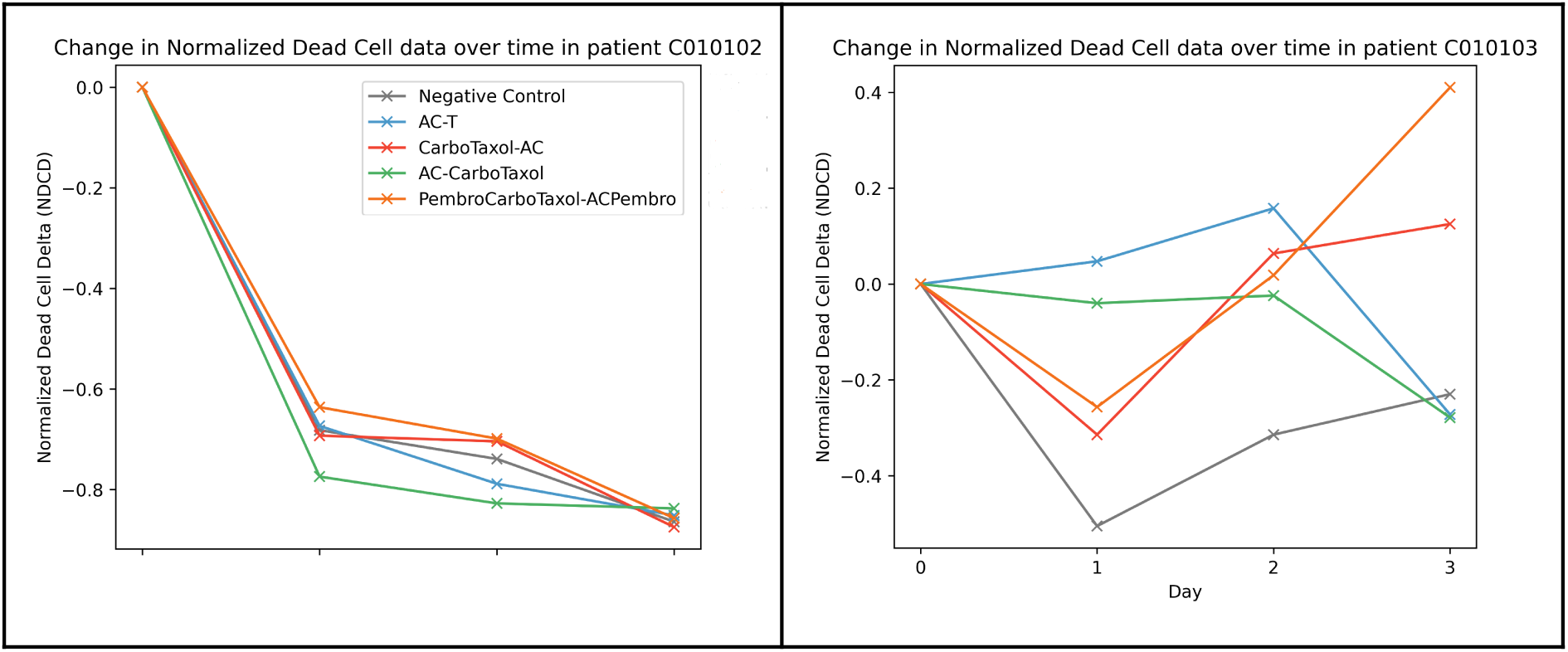
Changes in Normalized Dead Cell Delta (NDCD) over time in two patients; higher values denote a greater amount of cell death. A substantial difference in response to treatment between patients was observed. In the left-hand patient, none of the therapies have much effect over the negative control, suggesting that none of the commonly used regimens are likely to lead to pCR; in the right-hand graph there is a clear difference between the response to PembroCarboTaxol-ACPembro (ie. neoadjuvant Keynote-522 regimen) and the negative control, but also differential response to different agents. The complete data for all patients can be seen in Appendix 4.2.

On an individual patient basis, there were significant differences between responses to the same neoadjuvant therapy regimens between patients (Figure 3; App4.2). Mean Kendall’s W for the 12 patients was 0.16 denoting poor correlation between metrics. This indicates unique information that can be extracted from each metric, as well as the general lack of superiority or inferiority of any given treatment regimen at a population-level. (Figure App4.3).

Across the population, there was a significant difference in the rank-order of effective therapies: In 5 patients, AC-CarboTaxol had the greatest impact; in 3, PembroCarboTaxol-ACPembro ; in 3 CarboTaxol-AC; in 1, AC-T had the greatest response (Figure 4). No treatment regimen demonstrated consistent superiority or inferiority across patients.

**Figure 4:**
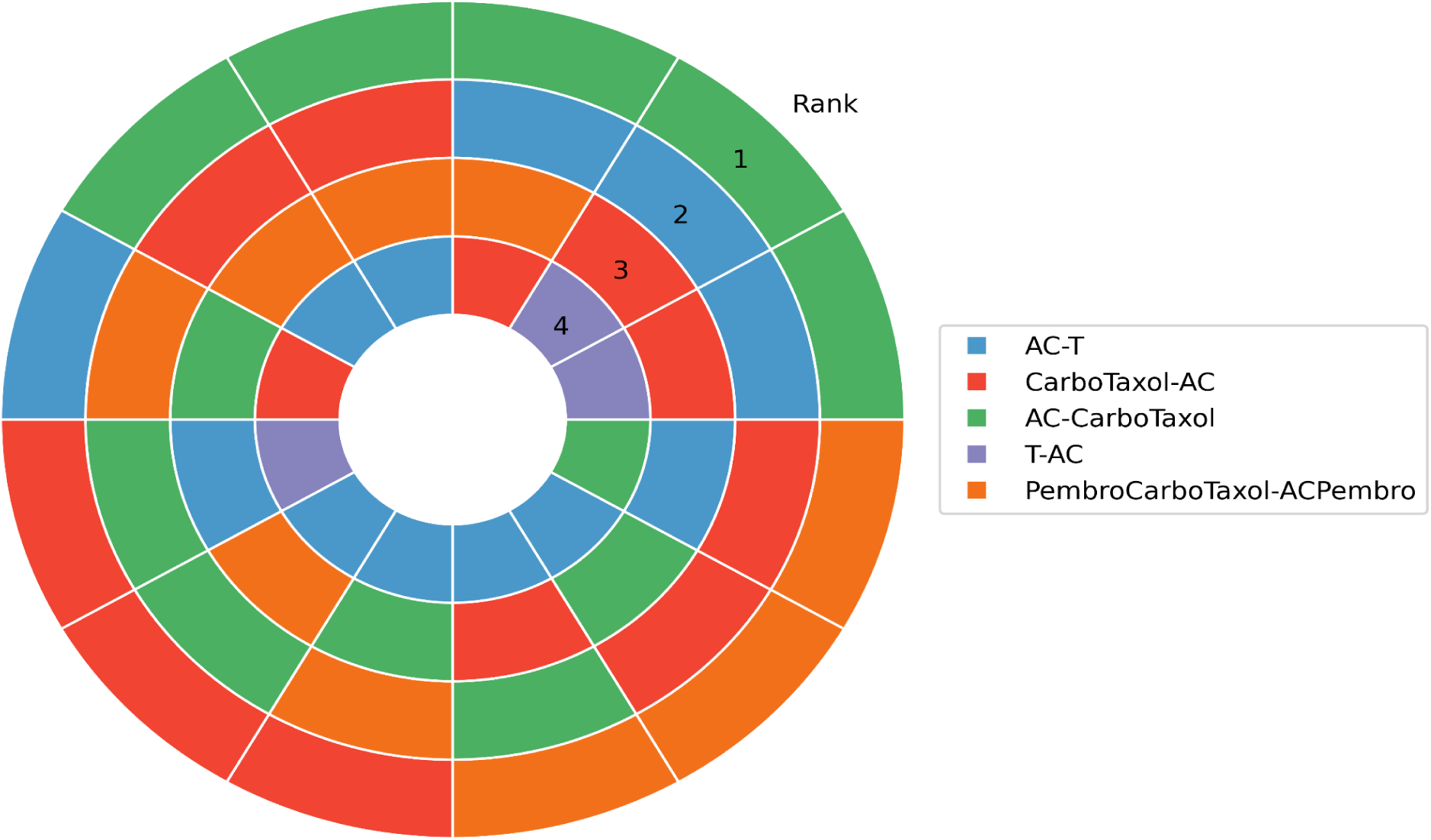
Stacked donut plot showing the ranked order of predicted effectiveness of each treatment regimen per evaluable patient. The outermost ring is the most effective treatment, the innermost ring is the least effective treatment. Each patient has treatment effectiveness ranked individually such that each patient is one slice, with a total of 12 slices (12 evaluable patients) to compare rankings between patients.

### Predicting pCR

Our analytical cohort consisted of 12 patients (9 pCR; 3 non-pCR). We generated ROC curves for each metric, and used those to identify the optimal cutoff for each metric and assessed performance by calculating a confusion matrix and a separation plot. The test predicted pCR well: All 7 of those predicted to achieve a pCR did achieve one. Specificity was 100% and sensitivity 78%, with an accuracy of 83%. Positive Predictive Value was 100%, and Negative Predicitve value of 60%. The p-value for the confusion matrix was 0.0455.

In the 3 patients who did not achieve a pCR, we explored whether there were other regimens that achieved a better response *ex vivo*, as measured by NDCD, than the regimen given. For two patients, there was no other regimen tested that was better than the one delivered. One patient received the Keynote-522 regimen, where AC-CarboTaxol was predicted to lead to a better response (Figure 6c). Using the cut-off established from the ROC curve (Figure 5), the response to Keynote-522 was below the value that would predict pCR, and the response seen with AC-CarboTaxol was above the threshold value that would predict pCR.

**Figure 5:**
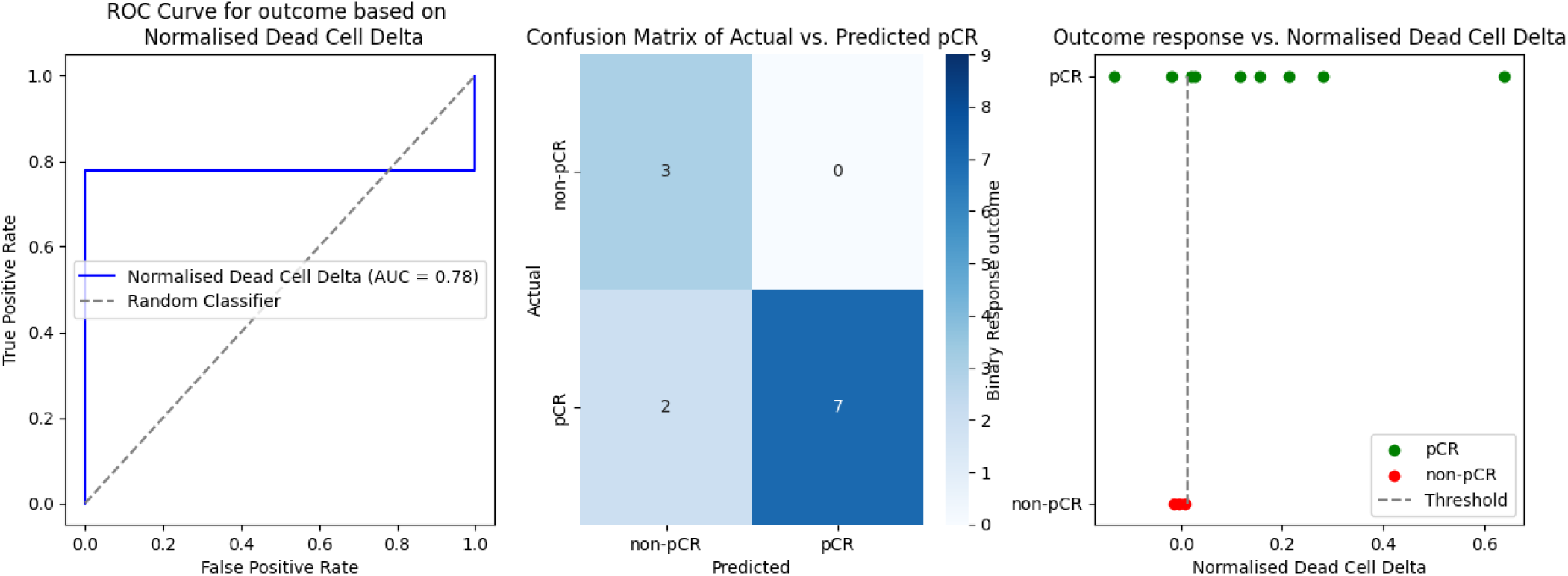
ROC curve, Confusion matrix and Separation plot for the 12 evaluable patients. Specificity 100%, Sensitivity 78%, Accuracy 83%, p-value = 0.0455.

**Figure 6:**
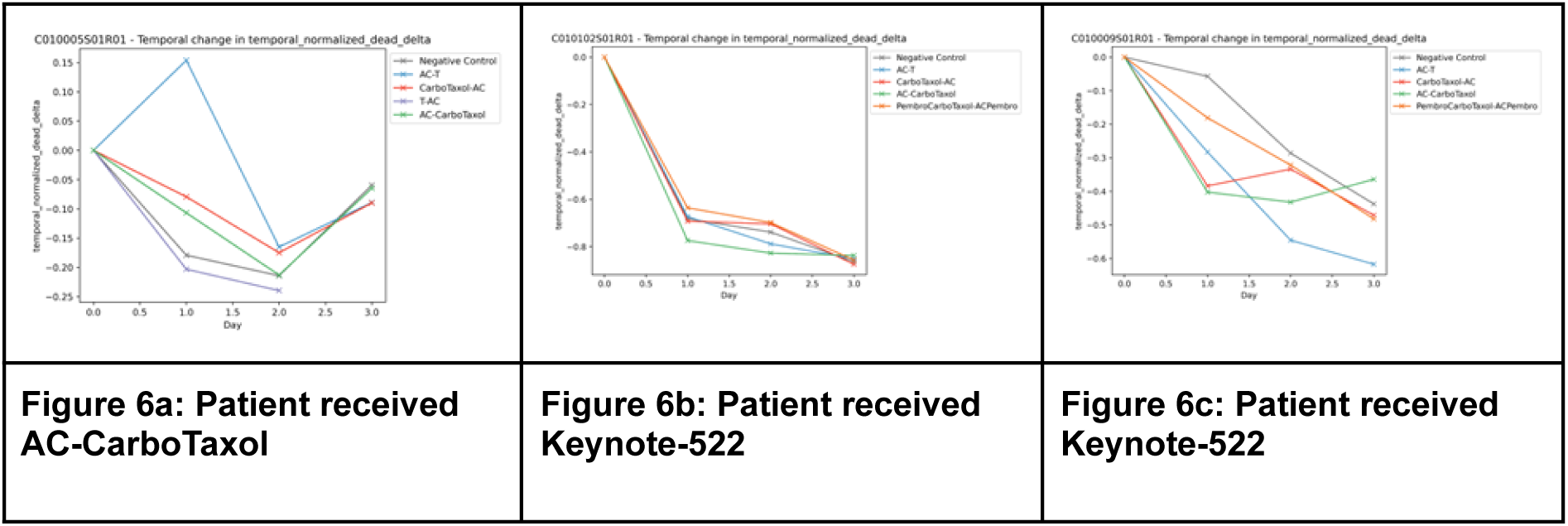
Per-patient response in the three patients where treatment did not lead to a pCR. X axis is days. Y axis is normalized dead cell delta (NDCD): higher values denote a greater response to treatment. 6a and 6b show that no other tested regimen shows a better in-assay response that the regimen received. 6c shows that in-assay response is better with at least one other regimen than the one delivered.

## Discussion

We have presented the interim results of a clinical trial of a novel functional precision medicine assay in patients with newly diagnosed localized triple-negative breast cancer undergoing neoadjuvant therapy (Cohort A of PEAR-TNBC). Patients underwent treatment as per standard of care, and we compared predicted response to actual response. The trial enrolled 34 patients, of whom 12 were evaluable at the time of analysis (31^st^ July 2024).

The median age was 53 years. 5 patients were White, and 4 Asian, and the most common staging was T2N0 (stage IIA). The most common regimens were Keynote-522 and AC-CarboTaxol. 9 out of 12 patients (75%) achieved a pCR. Of the 3 patients that did not achieve pCR, 1 patient had a regimen identified (AC-CarboTaxol) that was predicted to perform better than the regimen delivered (Keynote-522); in addition, the in-assay response to AC-CarboTaxol exceeded the threshold value of response that would predict pCR. It therefore suggests that, had the patient received a different treatment, they may have achieved a pCR, despite receiving a regimen that has been shown in an RCT to perform better at a population level.

It is important to note that these are interim data, and as the trial data matures, the exact cut-offs and performance may vary. In particular, we expect the pCR rate to drop as it approaches the long-term expected value of 55-60%. Nonetheless, this data clearly indicates feasibility and early support for reasonable performance in predicting pCR.

Previous work on predicting response to treatment in TNBC is limited. Apart from tumor size, there are a range of different potential biomarkers, including tumor infiltrating lymphocytes (TiL), Ki-67, multi-gene homologous recombination signatures and BRCA-1 status for PARP inhibitor response in the adjuvant setting (9,20,21). However, with the exception of BRCA-1, these are predictors of response to treatment in general, rather than predicting response to different agents or regimens. Improving pCR rates is a major focus of early-stage breast cancer trials. Large programs such as I-SPY2 have increased the pCR rate through a combination of both extensive testing and access to new drugs.

Clinical trials of FPM approaches are limited. Although there have been several approaches, most have very limited validation. One trial of 78 patients with relapsed glioma showed that using assay-directed therapy improved median overall survival from 9 to 12.5 months (6). Other work is mainly focused in hematological malignancies (22) and most previous work on FPM has been limited by the inability to include the impact of immunotherapy-based drugs. There is a useful review by Letai et al. that provides a good overview of the field (23). One additional regulatory hurdle is whether such approaches should be licensed as (in FDA terms) Companion or Complementary Diagnostics (24,25).

The main benefit of functional assays is that they directly measure the impact of therapy. Clinically, this provides a route to predicting the impact of therapy in patients with cancers that lack biomarkers, but also the potential to compare the impact of different therapies in a head-to-head manner, and to compare the impact of combinations against single agent therapy. This is in contrast to existing precision medicine approaches that identify genetic abnormalities and then associate those with a likely response to treatment, but do not measure it directly. However, this needs to be balanced against the considerable challenges around development. One of the main challenges is the need for fresh or viably cryopreserved tissue, not just for analysis, but also for assay development, and the inevitable problems with quality assurance and reproducibility that come from using fresh or cryopreserved human material. In addition, practical problems around access to tissue may limit its use in some indications (e.g. pancreatic cancer or brain tumors) where accessing fresh tumor tissue can be difficult. There are other potential uses of such functional assays, including drug development and target discovery, but these lie outside the scope of this report.

Our analysis of performance is based on a single-metric based prediction of pCR. Given the small patient numbers, this is appropriate. However, the assay and CV pipeline makes it simple to produce a range of metrics. Therefore, as patient numbers accrue, we will explore the use of more complex metrics (e.g. measures of migration speed) and the integration of multiple metrics, including the use of machine learning models. In addition, manual review of the imaging has shown issues with the dyes used: MitoView staining can persist even after cell death, and NucView only stains cells dying via the caspase-3/7 route, rather than other routes to cell death. As a result, Cohort B will use an improved dye system, and will co-incubate tumor-resident cells with peripheral blood mononucleocytes (PBMCs) from a blood sample that each patient will provide.

The focus of this paper is on assessing the ability of a novel functional precision medicine assay to predict pCR in newly diagnosed TNBC patients. The intention is that patients with a high risk of not achieving pCR could be offered access to different therapies, including clinical trials. It is notable that in those 3 patients who did not achieve pCR with the therapy they received, 1 patient had a regimen that was more highly ranked than the regimen received. Such an assay therefore has the potential to be both prognostic (i.e. predict pCR vs. non-pCR) but also predictive (i.e. predict which drugs may be most effective), although this clearly requires further validation. Such assays also open up the route to further research, helping understand the biological basis of resistance to treatment in patients who do not achieve pCR, and provide a test-bed for target and drug discovery and early development of new agents. The trial continues to collect outcome data on Cohort A and to enroll patients in Cohort B.

## Data Availability

Data are not publicly available due to the sensitive commercial nature of the data

## Acknowledgements

Mention trial sites and nurses

- BioRender for illustrations
- Arden Tissue Biobank for tumor tissues involved in optimization
- Dr. Gastón Primo for hydrogel optimization
- Dr. Marko Storch at Biofoundry (I-Hub) and Imperial College BRC Genomics Facility for assistance with RNAseq experiment design and execution.

## APPENDICES

## Appendix 1: Drug supply sources and concentrations used

Doxorubicin (MedchemExpress, HY-15142): 1.94µM

Cyclophosphamide (MedchemExpress, HY-17420): 2.345mM Paclitaxel (MedchemExpress, HY-B0015): 23nM

Carboplatin (MedchemExpress, HY-17393): 539µM

Pembrolizumab [Keytruda] (Merck (via everyone.org, CAT number): 25 ug/mL

## Appendix 2: Pre-clinical optimization of drug doses

**Figure App2.1.**
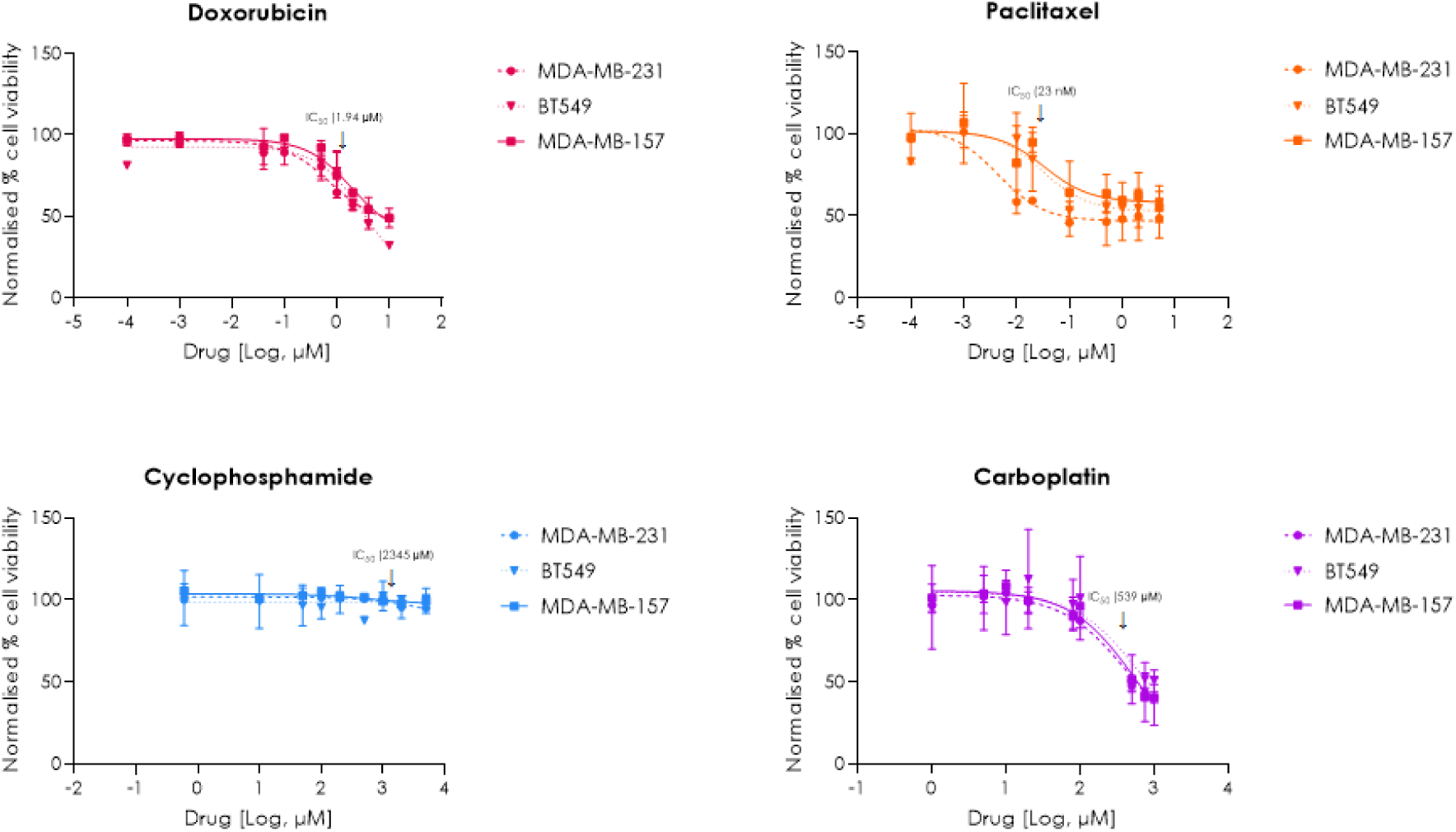
TNBC cell lines MDA-MB-231, BT549 and MDA-MB-157 were treated with chemotherapy drugs at indicated concentrations and imaged daily using an Incucyte SX5 system. Dose-response curves represent normalized confluency compared to DMSO/H2O as measured using the Incucyte at 24h for doxorubicin and cyclophosphamide, and 48h for paclitaxel and carboplatin (n=3).

**Figure App2.2.**
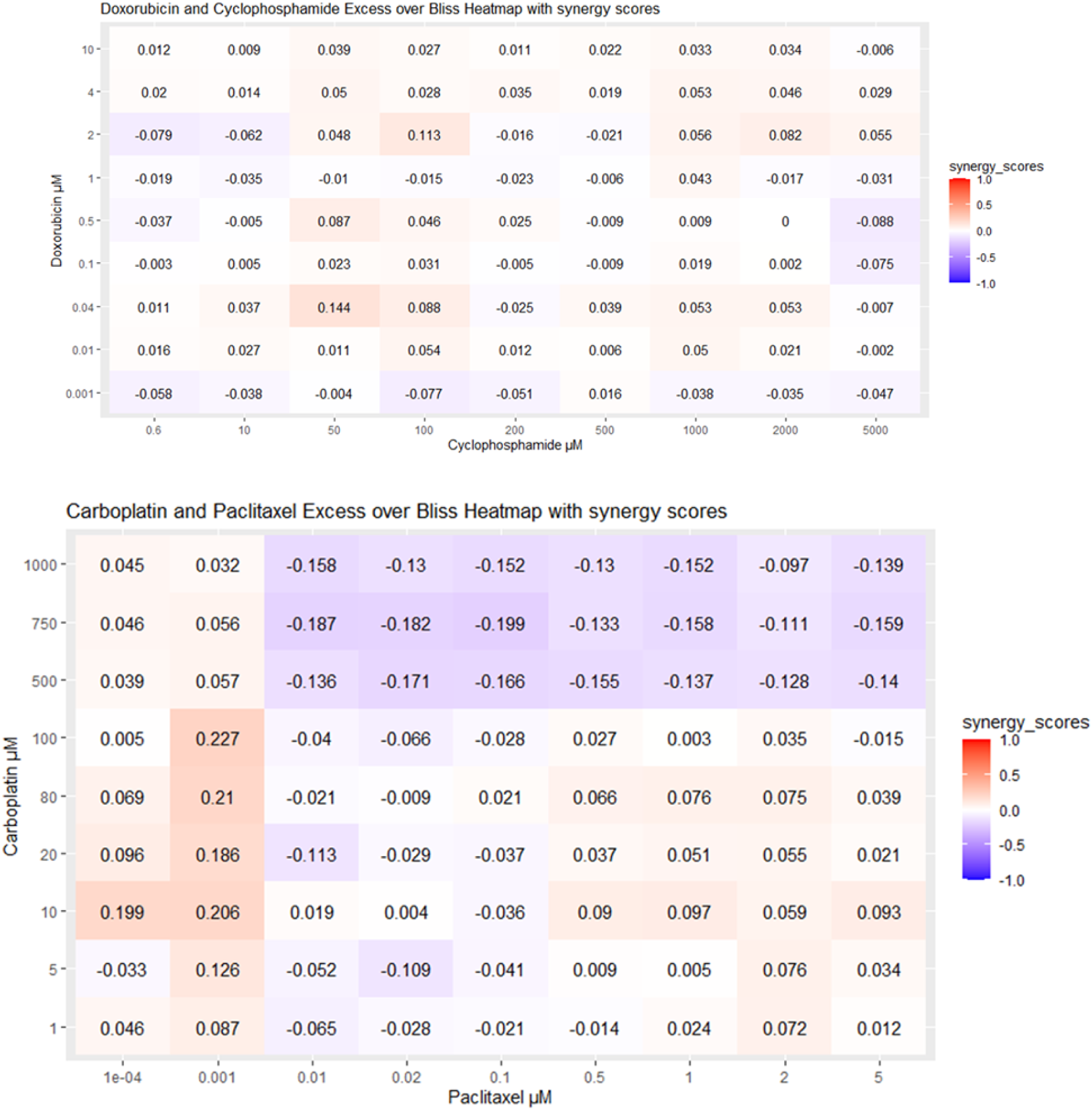
Excess over Bliss scores for MDA-MB-231 cells treated with Doxorubicin and Cyclophosphamide or Carboplatin and Paclitaxel in combination at indicated concentrations. Scores >0 indicate synergy, 0 = additive effect and <0 indicates antagonism (n=3).

**Fig App 2.3:**
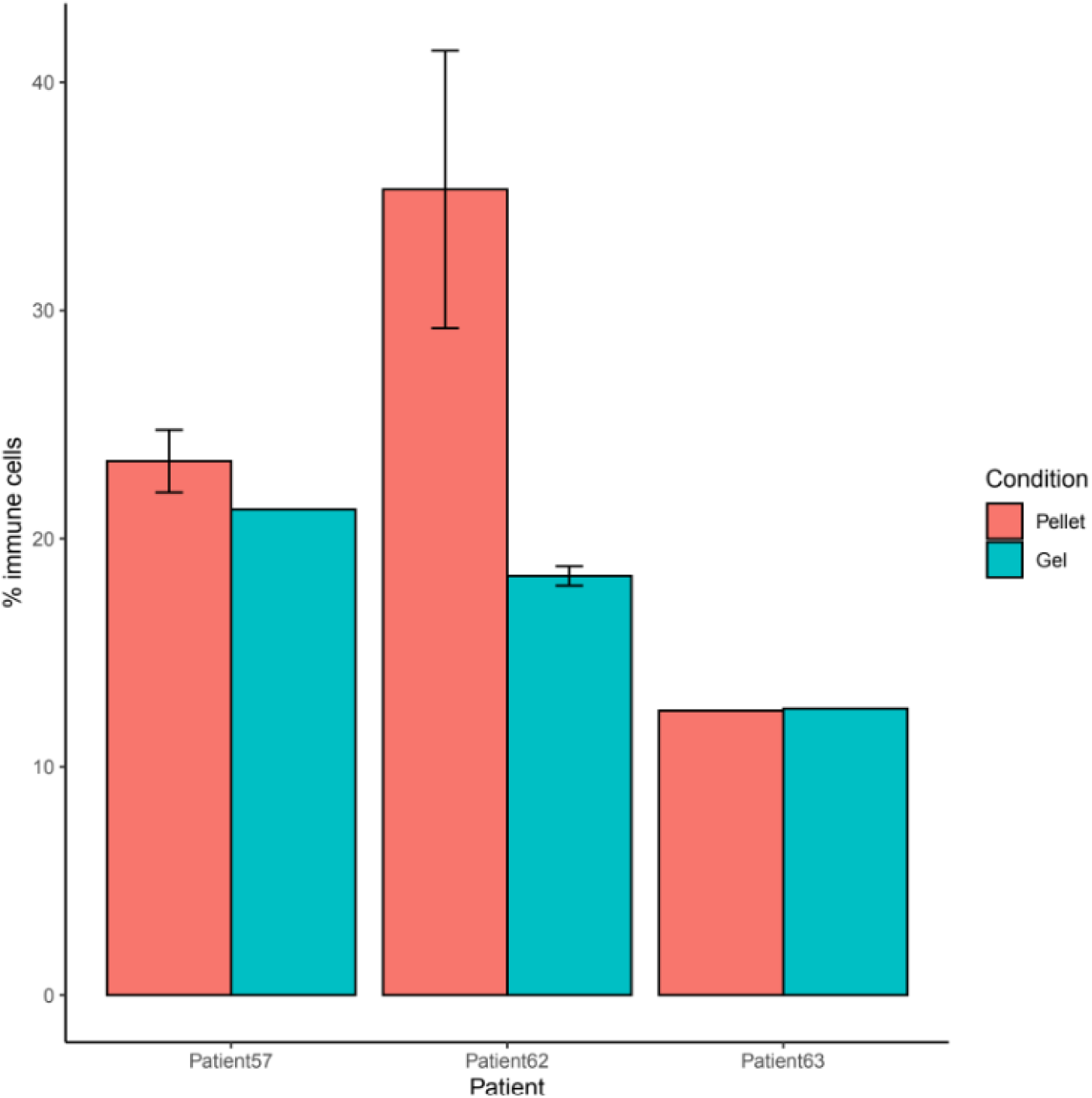
RNA-Seq cell deconvolution from 3 breast cancer patient samples. Tumor samples were processed for single cell extraction. Post extraction, cells were split for RNA extraction directly from pellets (“Pellet” condition) or 3D cell culture in BREA-F49 for 4 days (“Gel” condition). Cell deconvolution shows tissue resident immune cells are retained during the cell extraction process and that their percentage is further conserved upon 4 days of culture in 3D cultures (N=3).

## Appendix 3: Trial sample data and workflow and device images

**Figure App3.1:**
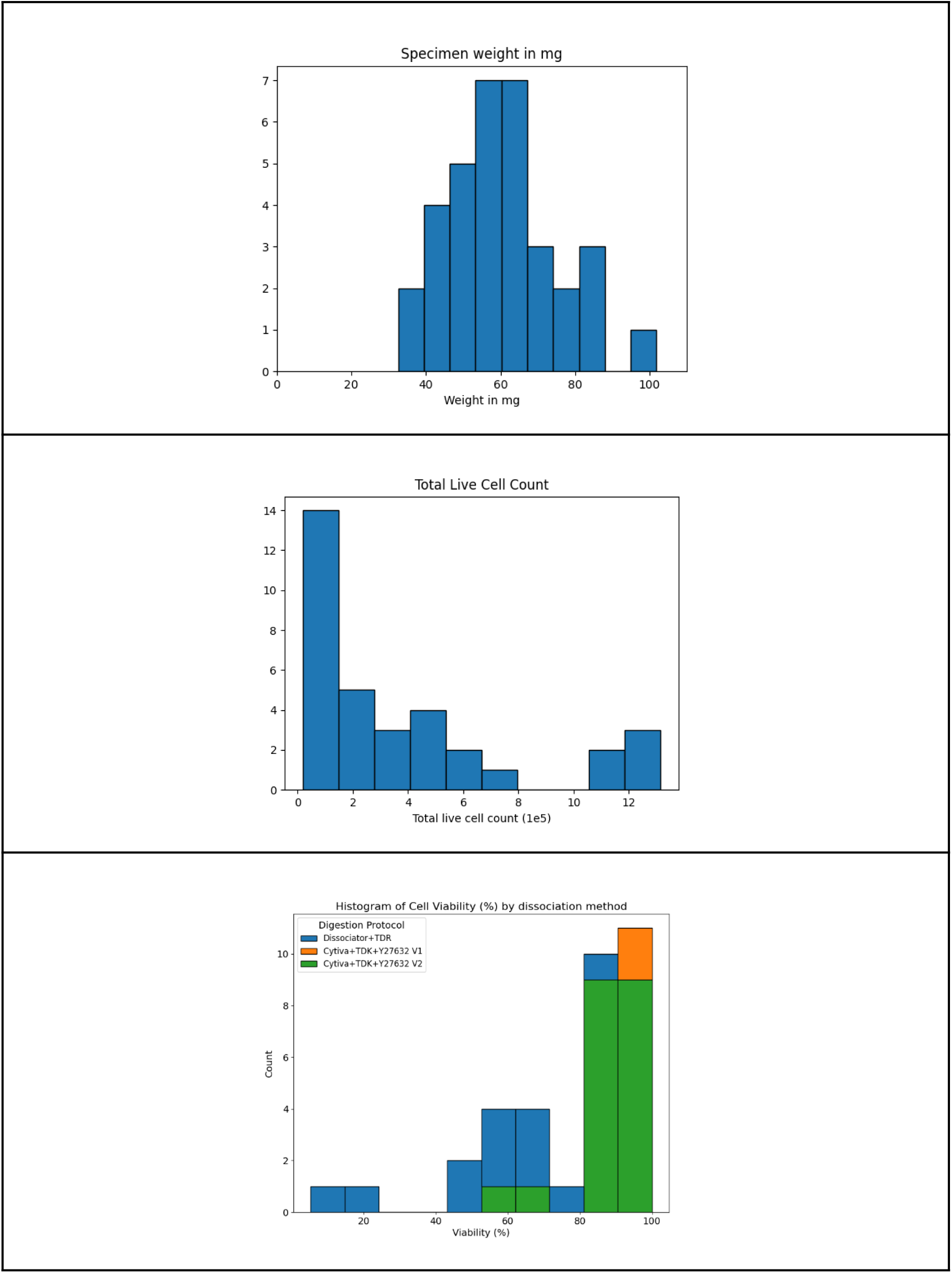
Sample weights, absolute number of live cells, and cell viability for patients in PEAR-TNBC (n=34)

**Fig App3.2:**
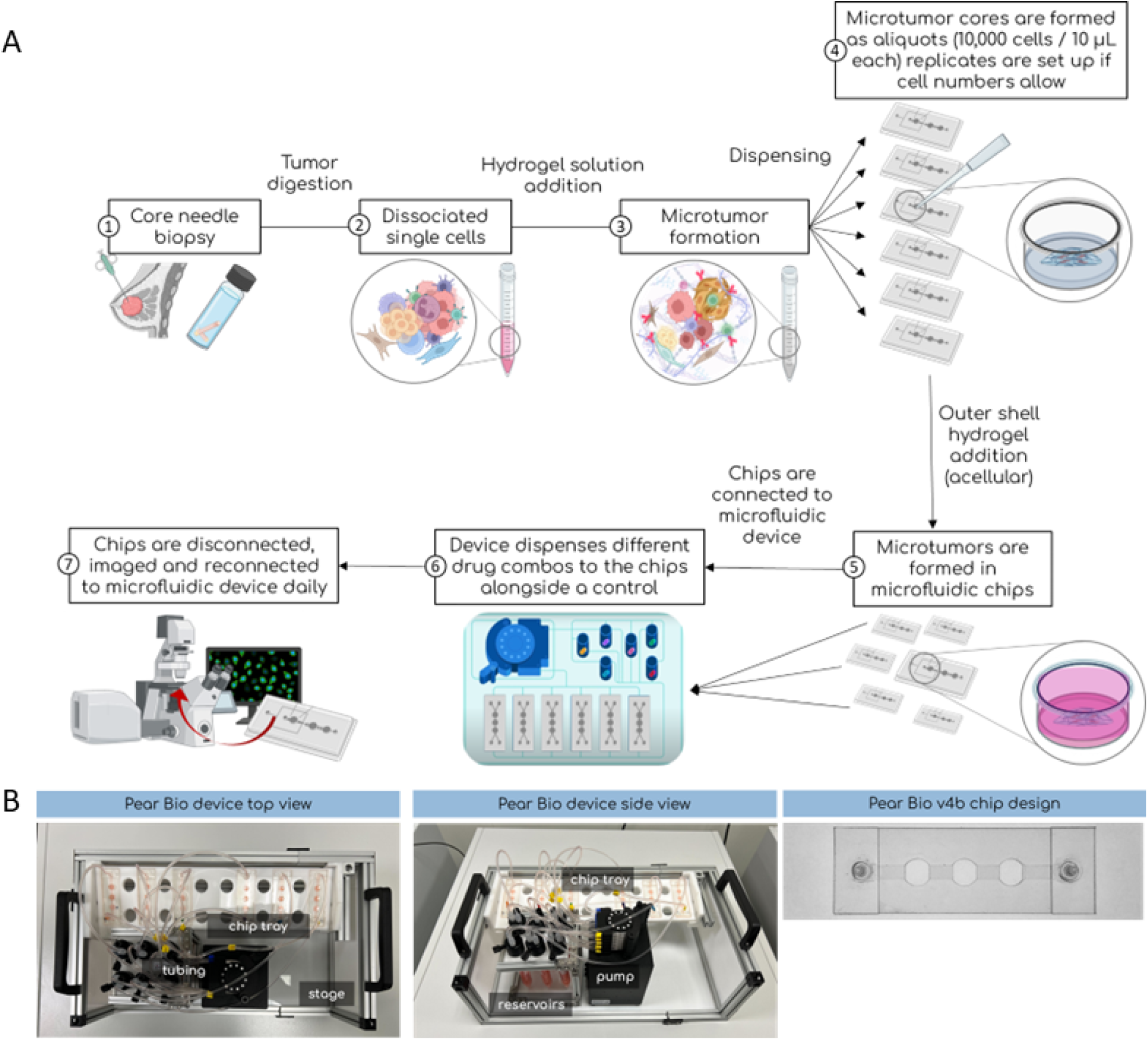
Process from core needle biopsy to testing (A) and images of the device and chip used (B)

## Appendix 4: Per-patient assay results

**Figure App4.1:**
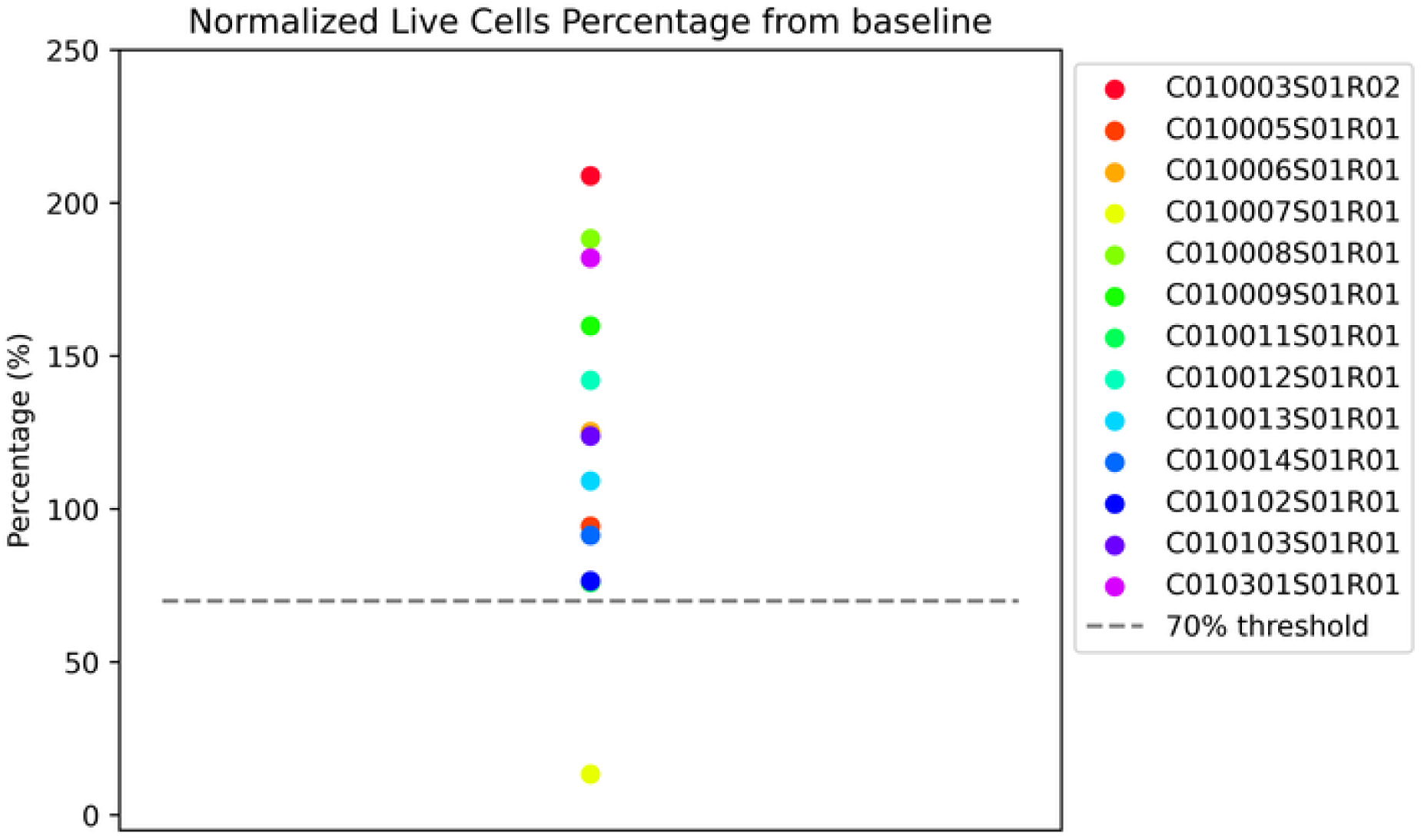
Change in Normalized Live Cell counts from baseline in the Controls chips. n=13: 12 evaluable patient, plus 1 patient where viability was not maintained in the control chip (as indicated in Fig 2 CONSORT Diagram).

### Appendix 4.2

**Figure App4.2:**
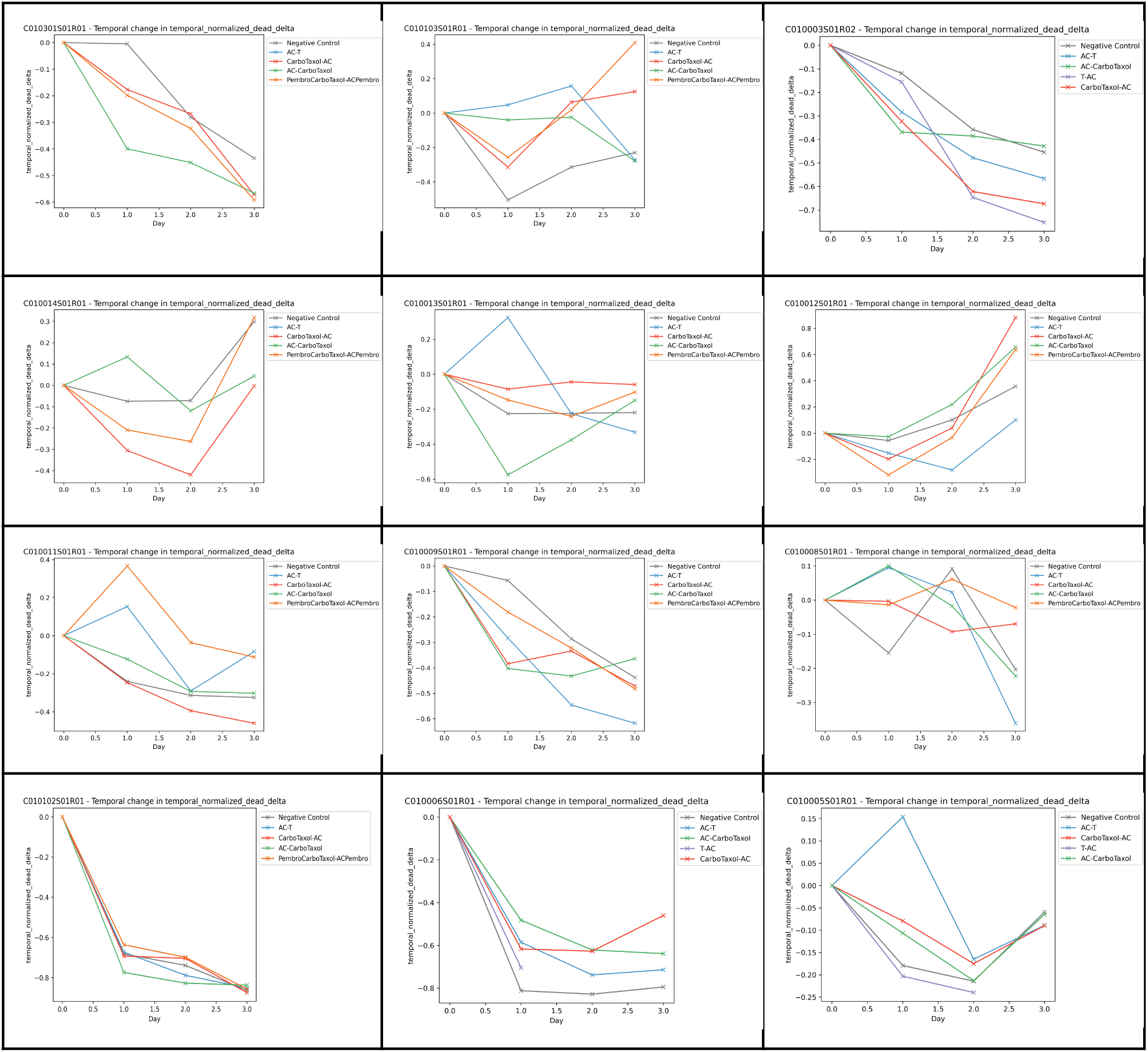
Per-patient assay results for Normalized Dead Cell Delta (NDCD) for all 12 evaluable patients in PEAR-TNBC. Each sub-graph is a separate patient; each line on the graph is a single microtumor treatment condition. Higher values are a greater change in dead-cell, and thus represent more cells dying.

**Figure App4.3:**
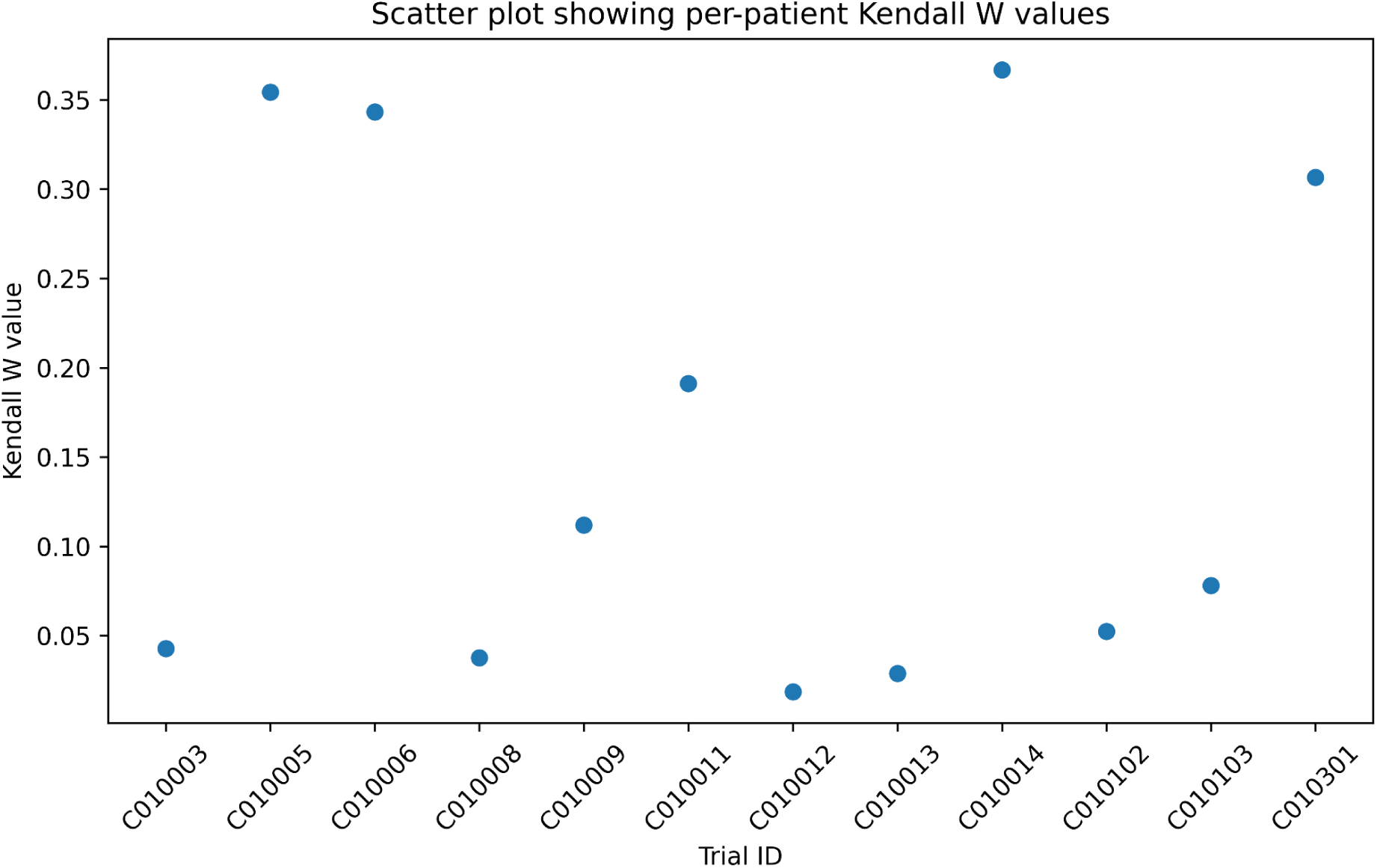
Kendall’s W values for each patient. Higher values indicate higher correlation between the different assay metrics. Although values vary between patients, the highest Kendall’s W was 0.36, denoting poor correlation between metrics. This indicates unique information that can be extracted from each metric, as well as the general lack of superiority or inferiority of any given treatment regimen at a population-level.

